# COVID-19 Outcomes in Patients with Cancer: Findings from the University of California Health System Database

**DOI:** 10.1101/2021.09.07.21263244

**Authors:** D. H. Kwon, J. E. Cadena Pico, S. Nguyen, K. H. R. Chan, B. Soper, A. L. Gryshuk, P. Ray, F. W. Huang

**Author notes:** **Corresponding authors** Dr. Franklin W. Huang, Division of Hematology/Oncology, Department of Medicine, University of California, San Francisco, 513 Parnassus Ave. HSE 1424, San Francisco, CA 94143, Dr. Daniel H. Kwon, Division of Hematology/Oncology, Department of Medicine, University of California, San Francisco, 505 Parnassus Ave. Rm M1286, San Francisco, CA 94143. **Twitter handles of authors** D. H. Kwon: @danielkwonmd, F. W. Huang @FranklingHuang.

## Abstract

**Background:** Patients with cancer are at risk for poor COVID-19 outcomes. We aimed to identify cancer-related risk factors for poor COVID-19 outcomes.

**Patients and Methods:** We conducted a retrospective cohort study using the University of California Health COVID Research Data Set. This database includes prospectively-collected, electronic health data of patients who underwent testing for SARS-CoV-2 at seventeen California medical centers. We identified adult patients tested for SARS-CoV-2 between February 1, 2020 and December 31, 2020, and selected a cohort of patients with cancer using diagnostic codes. We obtained demographic, comorbidity, laboratory, cancer type, and antineoplastic therapy data. The primary outcome was hospitalization within 30 days after first positive SARS-CoV-2 test. Secondary outcomes were SARS-CoV-2 positivity and composite endpoint for severe COVID-19 (intensive care, mechanical ventilation, or death within 30 days after first positive test). We used multivariable logistic regression to identify cancer-related factors associated with outcomes.

**Results:** We identified 409,462 patients undergoing SARS-CoV-2 testing. Of 49,918 patients with cancer, 1,781 (3.6%) tested positive. Patients with cancer were less likely to test positive (OR 0.69, 95%CI 0.66-0.73, P<0.001). BCR-ABL-negative myeloproliferative neoplasms (polycythemia vera, essential thrombocythemia, and primary myelofibrosis) (OR 2.51, 95%CI 1.29-4.89, *P*=0.007); venetoclax (OR 3.63, 95%CI 1.02-12.92, *P*=0.046); methotrexate (OR 3.65, 95%CI 1.17-11.37, *P*=0.026); Asian race (OR 1.92, 95%CI 1.23-2.98, *P*=0.004); and Hispanic/Latino ethnicity (OR 1.96, 95%CI 1.41-2.73, *P*<0.001) were associated with increased hospitalization risk. Among 388 hospitalized patients with cancer and COVID-19, cancer type and therapy type were not associated with severe COVID-19.

**Conclusions:** In this large, diverse cohort of Californians, cancer was not a risk factor for SARS-CoV-2 positivity. Patients with BCR/ABL-negative myeloproliferative neoplasm and patients receiving methotrexate or venetoclax may be at an increased risk of hospitalization following SARS-CoV-2 infection. Further mechanistic and comparative studies are needed to explain and confirm our findings.

## Introduction

To date, the severe acute respiratory syndrome coronavirus 2 (SARS-CoV-2) pandemic has caused over 4.5 million deaths worldwide.^1^ Patients with cancer have worse COVID-19 outcomes, including a 56% increased relative risk of intensive care unit admission and a 66% increased relative risk of all-cause mortality.^2^

Multiple cancer-related risk factors, such lung cancer type and chemotherapy use, have been identified.^2–10^ However, outcomes of patients with less common cancer types are unknown given modest sample sizes of prior studies, limiting statistical power. Additionally, the contribution of individual antineoplastic systemic therapies and regimens to risk of poor outcomes remains unclear. Elucidating indeterminate or unknown risk factors is useful in counseling and management of patients with cancer and COVID-19.

To address these outstanding questions, we leveraged a novel data set, the University of California Health COVID Research Data Set (UC CORDS).^11^ This data set includes electronic health data of all patients who underwent testing for SARS-CoV-2 at University of California (UC)-affiliated hospitals. We hypothesized that certain cancer types and systemic therapies are associated with worse COVID-19 outcomes.

## Methods

We conducted a retrospective cohort study of patients using UC CORDS v2.0.^11^ This limited data set includes prospectively-collected electronic health data of all patients who underwent quantitative reverse transcription polymerase chain reaction (RT-qPCR) testing for SARS-CoV-2 at 5 UC academic medical centers (Davis, Irvine, Los Angeles, San Diego, and San Francisco) and 12 affiliated California hospitals. UC CORDS is organized using the Observational Medical Outcome Partnership common data model, which contains diagnoses, medications, labs, and procedures associated with clinical encounters. Data are refreshed on a weekly basis.

### Cohort definitions

We first identified patients who received a SARS-CoV-2 RT-qPCR test between February 1, 2020 and December 31, 2020, and were ≥18-years-old at first test date. We then identified a cohort of patients with cancer, defined as ≥1 clinical encounter associated with a cancer ICD10-CM code within 1 year prior to the test date. For patients with a positive SARS-CoV-2 RT-qPCR test, the first positive date of was used; otherwise, the first negative test date was used. Patients with only basal and squamous cell cutaneous cancers were excluded given the extremely low morbidity and mortality of these cancers. Patients with other/unknown gender were excluded given few cases. For analysis of severe COVID-19 (defined below), a third cohort of hospitalized cancer patients with COVID-19 was selected to enrich for laboratory data and likelihood that poor outcomes are attributable to COVID-19. Supplementary Data S1 shows a flow diagram of these three cohorts.

### Independent variables

For demographic variables, we abstracted birth year, gender, race, and ethnicity. For clinical variables, we abstracted cancer type; comorbidities known to be associated with COVID-19 severity in patients with cancer (i.e., coronary artery disease, congestive heart failure, chronic kidney disease, chronic obstructive pulmonary disease, and asthma within 1 year prior to test date); and body mass index (BMI).^12^ Cancer types were categorized based on organ system (e.g., urinary tract included renal cell and urothelial carcinomas). Antineoplastic systemic therapy use from 60 days prior to test date to 30 days after was abstracted. Antineoplastic systemic therapies were categorized by antibody, chemotherapy, hormone therapy, immune-based therapy, tyrosine kinase inhibitor, other cytotoxic therapy, and other targeted therapy (Supplementary Data S2). We also abstracted laboratory data from 60 days prior to the SARS-CoV-2 test date to 30 days afterward.

### Dependent variables

Outcomes included SARS-CoV-2 positivity (at least 1 positive RT-qPCR test); hospitalization within 30 days after first positive test date; and a composite endpoint for severe COVID-19, defined as either intensive care unit (ICU) admission, need for mechanical ventilation, or death within 30 days after first positive test date.

### Statistical analysis

We first calculated the incidence of SARS-CoV-2 test positivity among all patients tested for SARS-CoV-2, regardless of cancer. Then, we created a multivariable logistic regression model to predict risk of test positivity among this cohort, which included age, gender, race, ethnicity, comorbidities, any cancer history, and receipt of any systemic therapy. To identify cancer-related risk factors for hospitalization in patients with cancer and COVID-19 (the primary outcome), we created two multivariable logistic regression models: one in which therapies were categorized and another in which individual therapies were delineated. Lastly, to evaluate risk of severe COVID-19, we restricted the cohort to only hospitalized patients with cancer and COVID-19. We created a multivariable logistic regression model with systemic therapies as categories and with the addition of laboratory tests as continuous variables. We included only tests with a missing rate <30%. Laboratory tests were included only in this latter analysis given the high rate of missing values in non-hospitalized patients. We did not incorporate individual therapies, as the number of patients associated with each individual medication was small in this cohort. Multiple imputation was used for imputation of missing laboratory values.^13^

Logistic regressions were visualized using forest plots with odds ratios and 95% confidence intervals. *P*-values <0.05 were considered significant. We did not correct for multiple comparisons given the exploratory nature of the study.^14^ The logistic regression models were implemented using the statsmodels module in the Python programming language (v3.8).^15^ The study protocol was reviewed and approved by both UCSF and Lawrence Livermore National Laboratory institutional review boards.

## Results

Overall, 24,177 of 409,462 (5.9%) patients undergoing SARS-CoV-2 testing tested positive for SARS-CoV-2. Of the 49,918 patients with a history of cancer, 1,781 (3.6%) tested positive. The mean age of SARS-CoV-2-positive patients with cancer was 59 years (SD=16); 950 (53%) were female; 939 (53%) were White; and 636 (36%) were Hispanic or Latino (Table 1). The most common cancer types were Multiple (*N*=293, 16%); Breast (*N*=241, 14%); and Prostate (*N*=122, 7%) per Table 1. Three hundred and twenty-four (18%) patients were on active therapy, of which chemotherapy was the most common (*N*=153, 9%). Individual therapies are listed in Supplementary Data S3.

**Table 1.**
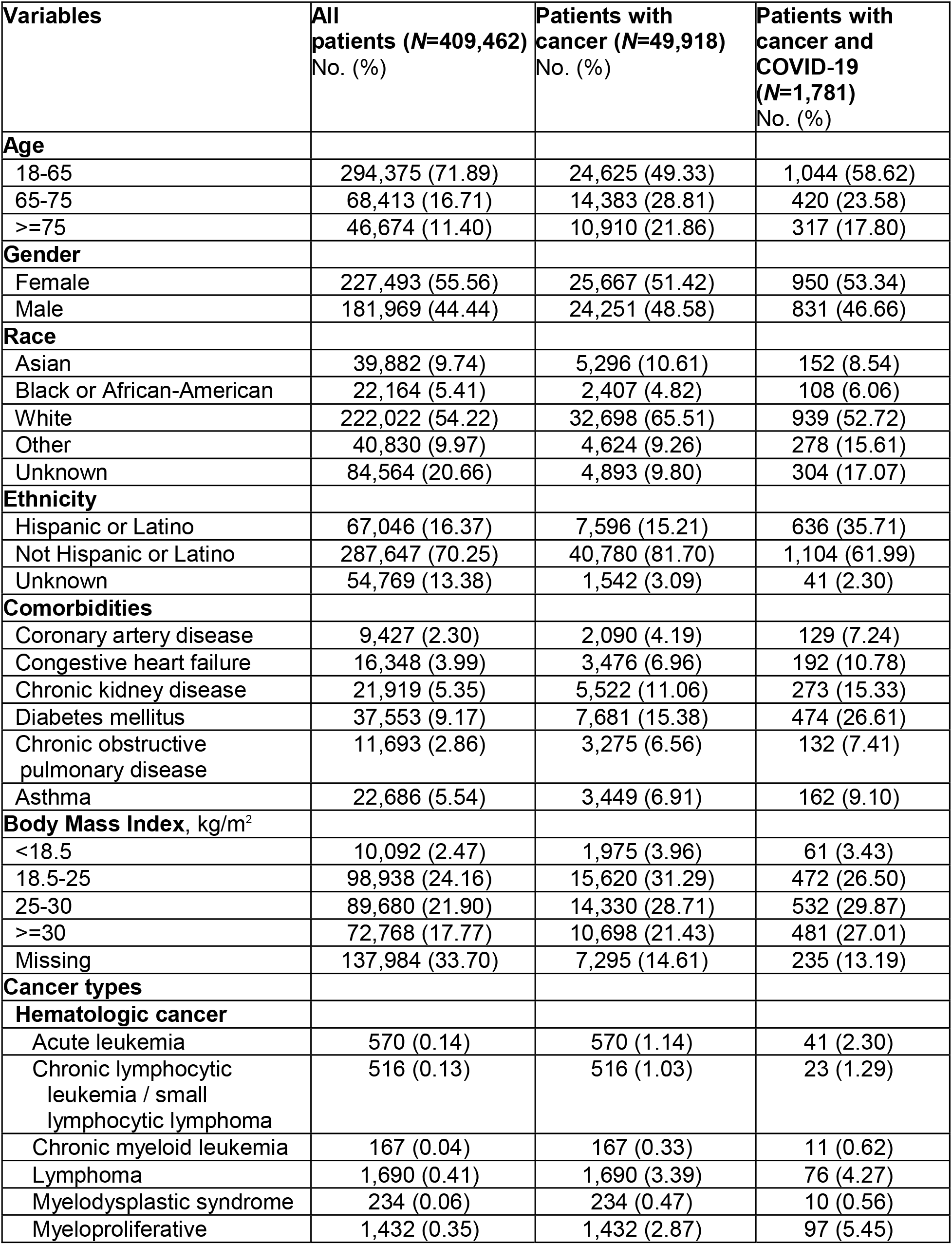
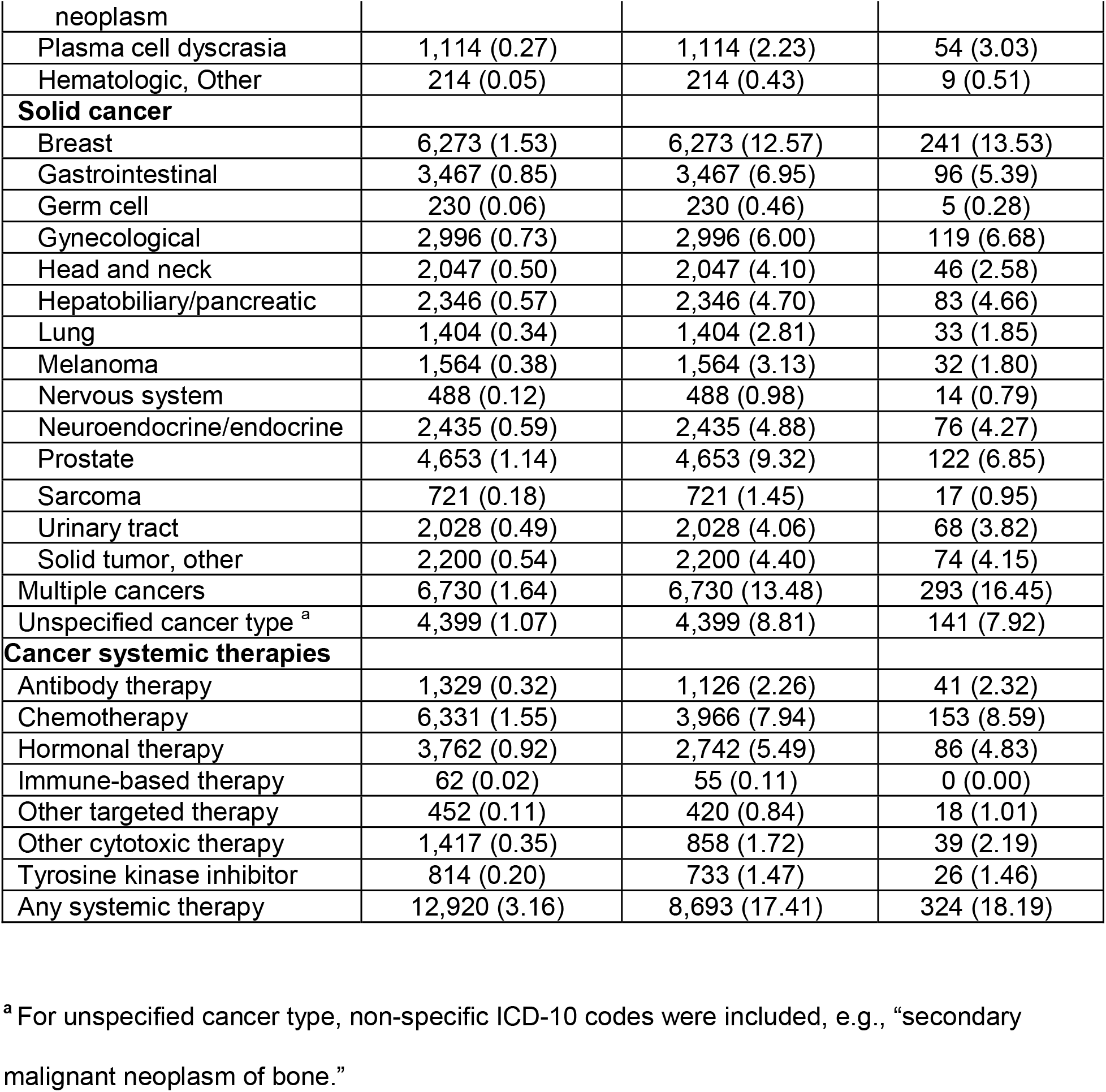
Characteristics of patients undergoing SARS-CoV-2 testing.

Figure 1 describes factors associated with SARS-CoV-2 test positivity in the entire cohort. In terms of cancer-related factors, positive cancer history was associated with a decreased risk of a positive test (OR 0.69, 95% CI 0.66-0.73, *P*<0.001). Many cancer types were associated with a decreased risk or no difference in risk of a positive test compared to unspecified cancer type (Supplementary Data S4). Similarly, any systemic therapy use was associated with a decreased risk of a positive test (OR 0.71, 95% CI 0.69-0.84, *P*<0.001).

**Figure 1:**
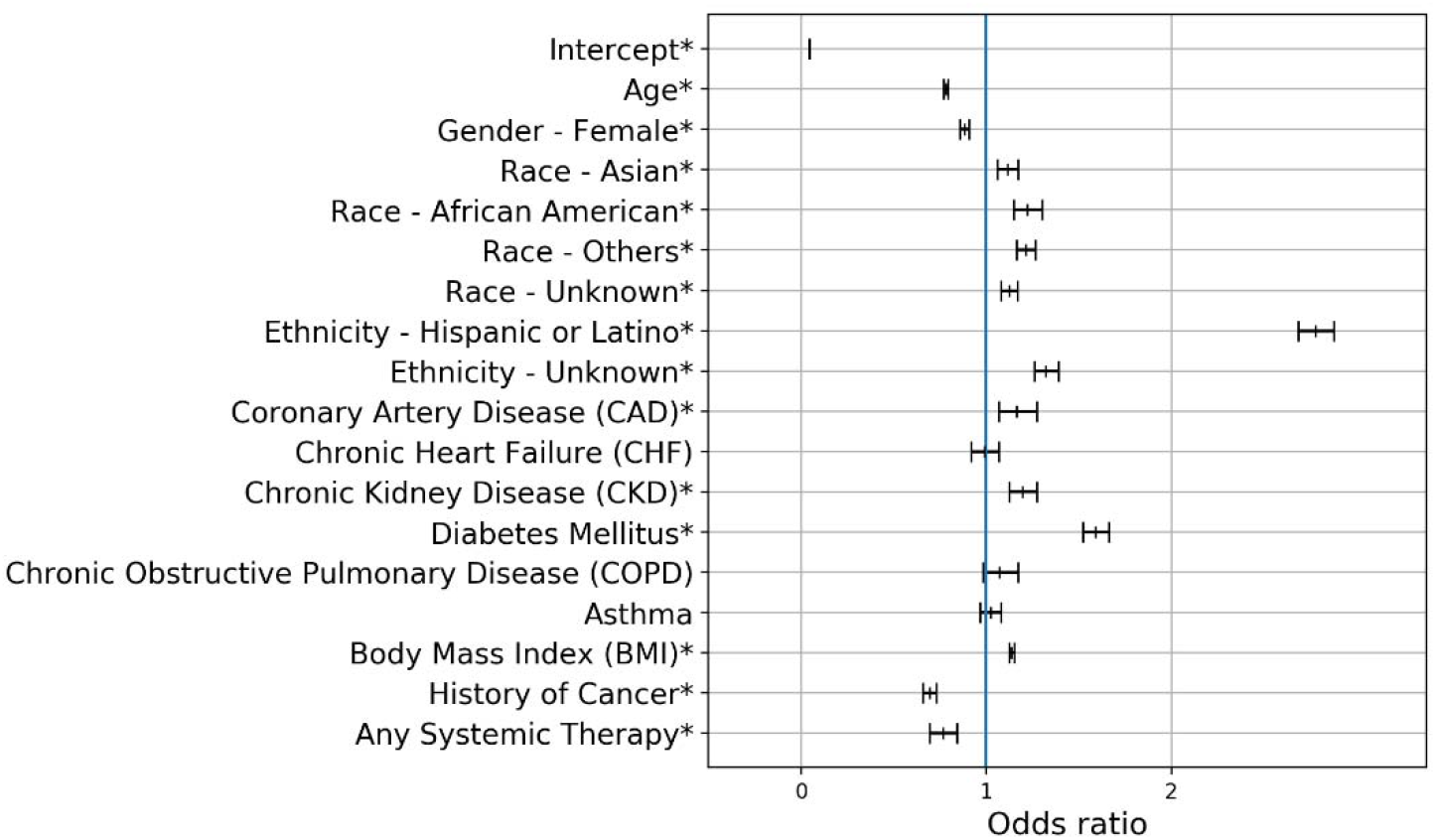
Risk of SARS-CoV-2 test positivity among 409,462 adult patients undergoing testing. In a multivariable logistic regression of all 409,462 adult patients who underwent SARS-CoV-2 testing, race, ethnicity, and comorbidities were associated with an increased risk of a positive test. History of cancer and antineoplastic systemic therapy were associated with a decreased risk of a positive test. Adjusted odds ratios are shown. Tabulated data can be found in Supplementary Data S6. * Denotes *P*<0.05

Among 1,781 patients with cancer, risk factors for hospitalization included older age, Asian race (compared to White), Hispanic/Latino ethnicity, and several comorbidities (i.e., coronary artery disease, chronic kidney disease, diabetes mellitus, and chronic obstructive pulmonary disease; Figure 2). In terms of cancer type, only myeloproliferative neoplasm (which includes polycythemia vera, essential thrombocythemia, and primary myelofibrosis and does not include chronic myeloid leukemia) was associated with an increased risk of hospitalization compared to unspecified cancer type (OR 2.51, 95% CI 1.29-4.89, *P*=0.007; Figure 2). For antineoplastic systemic therapies, chemotherapy was not associated with an increased risk of hospitalization (OR 1.46, 95% CI 0.95-2.26, *P*=0.086) and only Other Targeted Therapy was associated with an increased risk (OR 3.64, 95% CI 1.21-10.98, *P*=0.022). In the model with individual therapies, methotrexate (OR 3.65, 95% CI 1.71-11.37, *P*=0.026) and venetoclax (OR 3.63, 95% CI 1.02-12.92, *P*=0.046) were associated with an increased risk of hospitalization (Figure 3). Of note, venetoclax was categorized as an Other Targeted Therapy.

**Figure 2:**
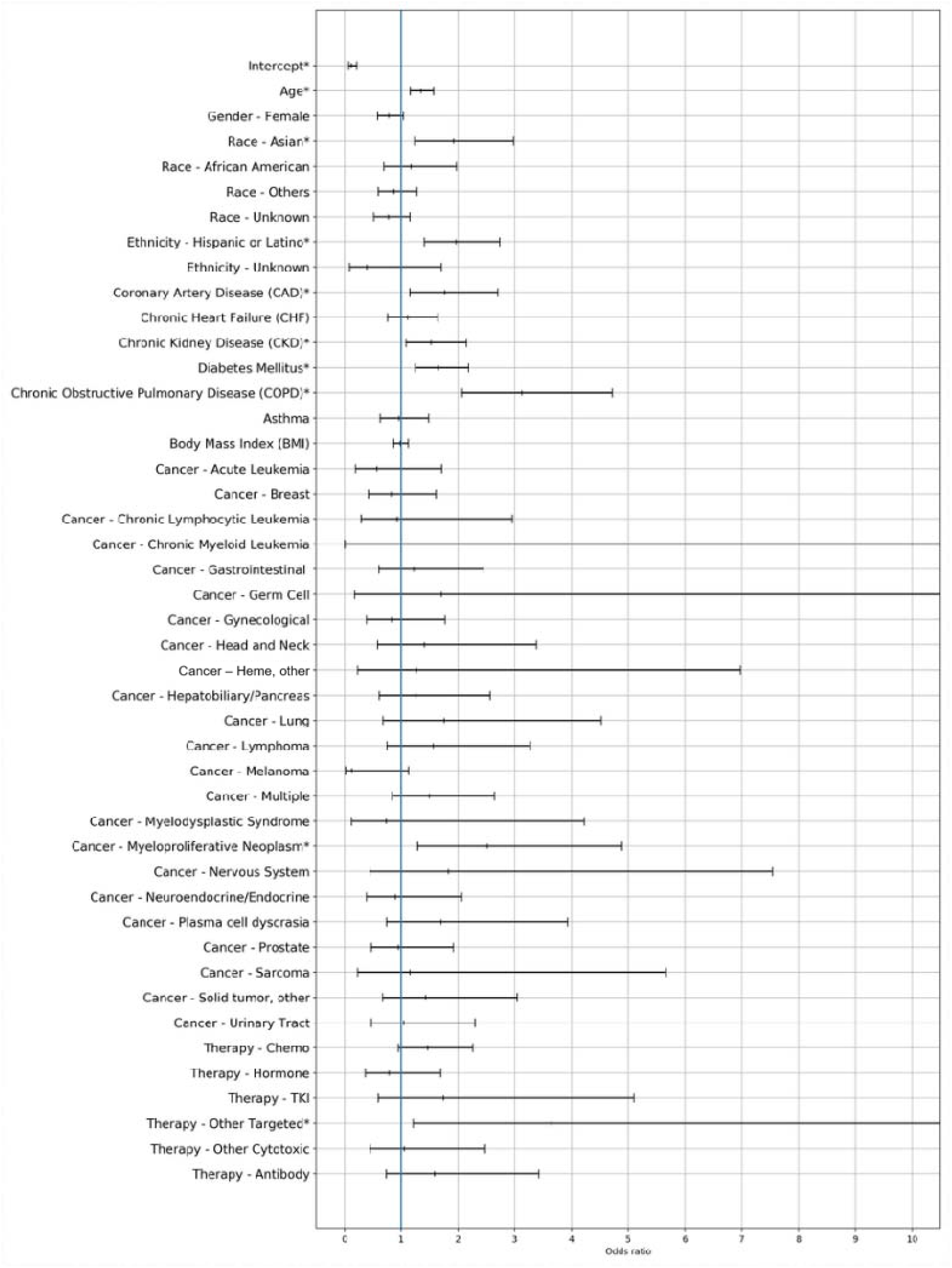
Risk of 30-day hospitalization following a positive SARS-CoV-2 test among 1,781 adult cancer patients. In a multivariable logistic regression of 1,781 adult patients with a history of cancer and a positive SARS-CoV-2 test, Asian race (compared to White race), Hispanic/Latino ethnicity (compared to not Hispanic/Latino ethnicity), multiple comorbidities, myeloproliferative neoplasm (compared to unspecified cancer type), and Other targeted therapy (compared to not) were associated with an increased risk of 30-day hospitalization. Adjusted odds ratios are shown. TKI = Tyrosine Kinase Inhibitor. Tabulated data can be found in Supplementary Data S7. * Denotes *P*<0.05

**Figure 3:**
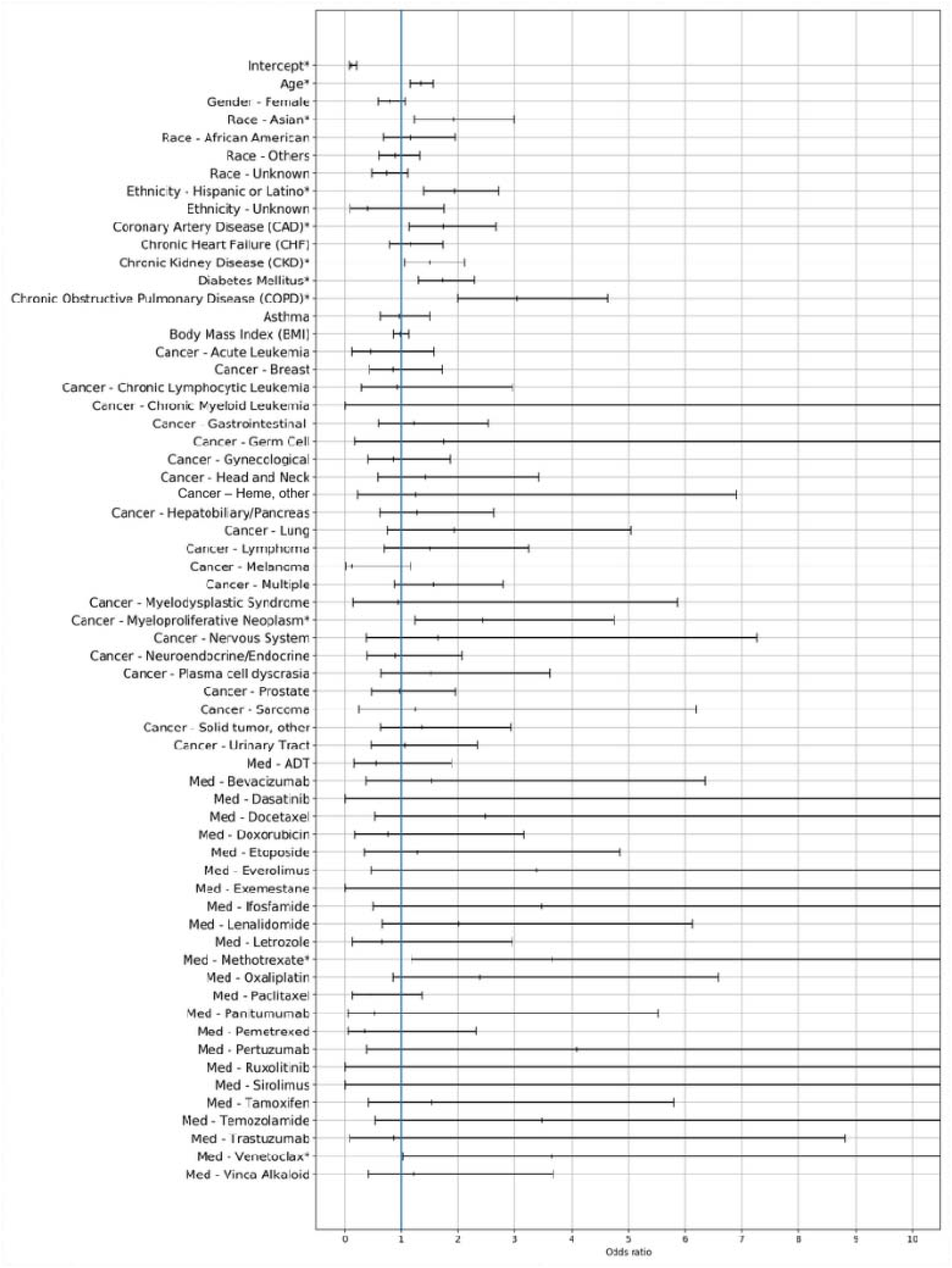
Risk of 30-day hospitalization following a positive SARS-CoV-2 test among 1,781 adult cancer patients, individual therapies delineated. This multivariable logistic regression is identical to that in Figure 2, except that individual systemic therapies are delineated. Methotrexate (categorized as chemotherapy) and venetoclax (categorized as Other targeted therapy) were associated with an increased risk of 30-day hospitalization. Adjusted odds ratios are shown. ADT = Androgen Deprivation Therapy. Tabulated data can be found in Supplementary Data S8. * Denotes *P*<0.05

In a *post-hoc* sensitivity analysis of this model, we changed the time window of therapy receipt from 60 days before through 30 days after positive test date to 60 days before through the positive test date. Methotrexate remained associated with increased risk of hospitalization (OR 4.12, 95% CI 1.10-15.46, *P=*0.036), but venetoclax did not (OR 3.63, 95% CI 0.91-14.53, *P*=0.068; data not shown). In another *post-hoc* analysis of systemic therapies and risk of positive SARS-CoV-2 test in the entire 409,462-patient cohort, venetoclax (but not methotrexate) was associated with an increased risk of a positive test (OR 2.35, 95% CI 1.39-3.97, *P*=0.001; data not shown).

Among 388 cancer patients who were hospitalized after a positive SARS-CoV-2 test, no demographic, comorbidity, or cancer-related factors were associated with a decrease in risk of severe COVID-19 (Figure 4). Higher albumin was associated with a decreased risk (OR 0.73, 95% CI 0.56-0.96, *P*=0.027), and higher glucose was associated with an increased risk (OR 1.44, 95% CI 1.10-1.90, *P*=0.008) of severe COVID-19. Myeloproliferative neoplasm (*N*=32) was not associated with severe COVID-19 (OR 0.70, 95% CI 0.20-2.48, *P*=0.582) compared to unspecified cancer type.

**Figure 4:**
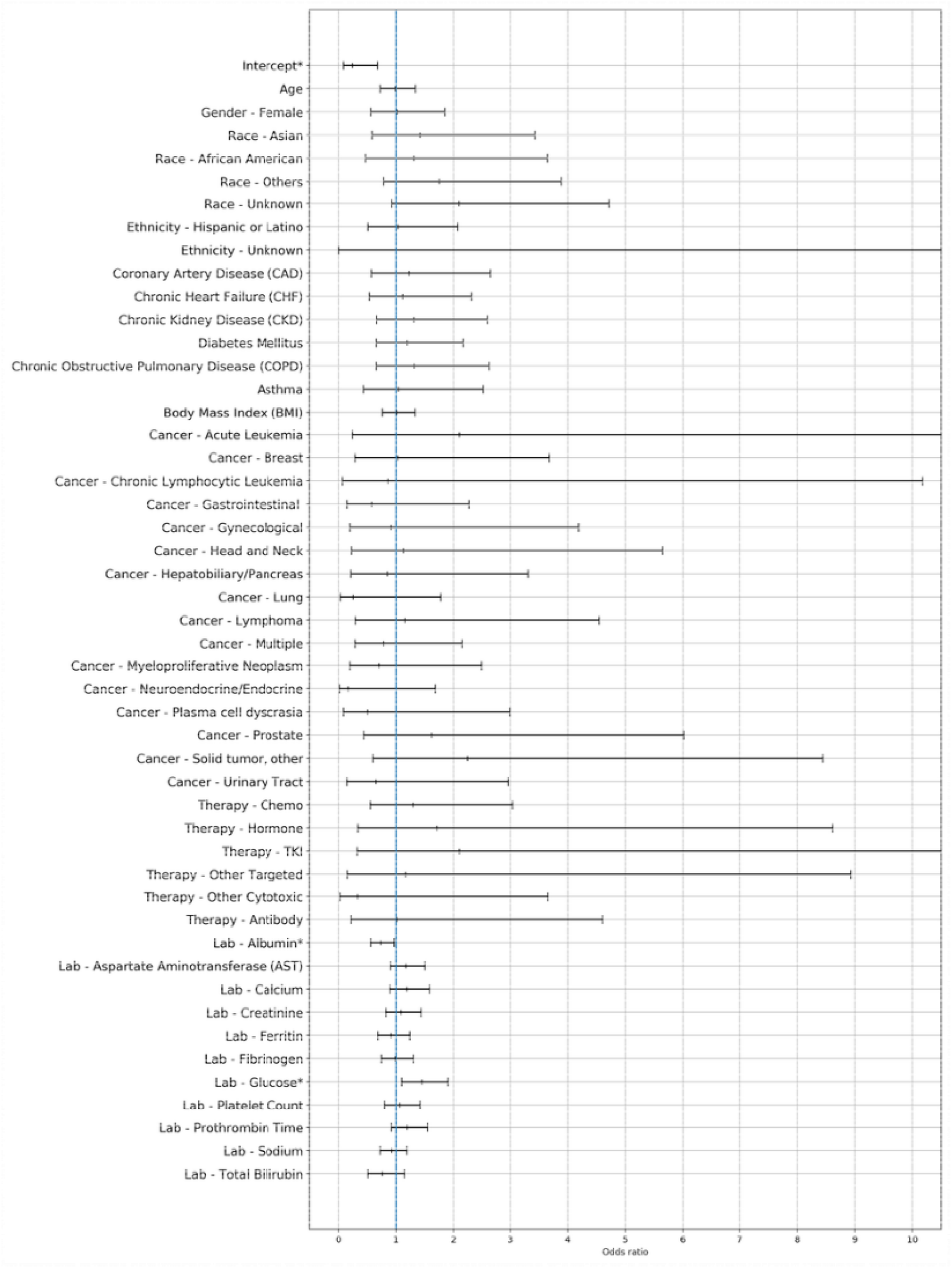
Risk of 30-day intensive care, mechanical ventilation, or death among 388 adult cancer patients with a history of cancer and positive SARS-CoV-2. In a multivariable logistic regression of 388 adult patients with a history of cancer who were hospitalized within 30 days after a positive SARS-CoV-2 test, no factors were associated with an increased risk of severe COVID-19. Severe COVID-19 was defined as intensive care admission, receipt of mechanical ventilation, or death within 30 days of first positive test. Adjusted odds ratios are shown. TKI = Tyrosine Kinase Inhibitor. Tabulated data can be found in Supplementary Data S9. * Denotes *P*<0.05

With the hypothesis that COVID-19 severity for patients with myeloproliferative neoplasms varies based on abnormalities in thrombosis-related laboratory values, we conducted a *post-hoc* analysis in which we added interaction terms between myeloproliferative neoplasms and platelet count and between myeloproliferative neoplasms and fibrinogen to the model. The interaction terms were not significant (Supplementary Data S5).

## Discussion

In this study, we used a novel multi-institution registry of passively and rapidly accumulating electronic patient-level data comprising all patients undergoing SARS-CoV-2 testing in UC health systems. We identified a large, diverse cohort of patients with cancer, and found new cancer-related factors associated with adverse outcomes.

In the COVID-19 and future pandemics, it is critical to rapidly identify patient-related attributes and interventions that affect the risk of infection, morbidity, and mortality. The timely creation of frequently and passively updated databases that contain patient-level clinical data from multiple health systems, like UC CORDS and the National COVID Cohort Collaborative (N3C), is invaluable to this goal. These data could complement those of other consortium and registry efforts, such as the COVID-19 and Cancer Consortium (CCC19), which provide more granular data using human abstraction.^4^ For example, Reznikov et al.^16^ mined UC CORDS within a few months of its creation and identified antihistamines associated with decreased SARS-CoV-2 test positivity. *In vitro* drug susceptibility assays showed hypothesis-generating antiviral mechanisms of candidate antihistamines.

Central data sets for future pandemics could include other data types (e.g., imaging, patient symptoms, and genomic data to identify mechanisms and therapeutic targets through techniques such as natural language processing); demographic data (e.g., patient location and activity using wearable devices); real-time data collection; and data from other health care systems (e.g., Veterans Health Administration and Community Health Centers). Pandemic preparation should also include identification of and resource-allocation to teams to harmonize and standardize raw data, and to query, analyze, and interpret databases. Efforts must be done ethically with potential biases in mind.^17^ Moreover, advanced analytic techniques, such as artificial intelligence (AI) approaches, that are findable, accessible, interoperable, and reusable (FAIR) to facilitate the development of new AI applications, could be applied.

In this study, we found a decreased risk of SARS-CoV-19 positivity in patients with cancer compared to those without cancer. This is counter to studies that report of an increased odds of infection in patients with cancer,^3,7,16^ and others that report no difference in risk.^18,19^ The discrepancy is unclear, but may be related to greater protective behaviors and testing practices in UC patients with cancer. For example, UC patients with cancer may be likely to employ behaviors that decrease transmission (e.g., social distancing and mask-wearing) or carry a lower threshold to undergo testing compared to patients with cancer in other regions. UC patients with cancer may also have been more likely to receive asymptomatic screening prior to infusions, radiation therapy, and surgeries, as had been instituted in some UC medical centers such as UCSF. We found that patients receiving systemic therapy were also less likely to test positive, perhaps for similar reasons. Future studies could examine such behaviors and SARS-CoV-2 testing indication to better investigate this discrepancy.

We also identified new risk factors for hospitalization in patients with cancer. Patients with BCR/ABL-negative myeloproliferative neoplasms and COVID-19 were at an increased risk of hospitalization. Patients with myeloproliferative neoplasms are in a pro-inflammatory state, with qualitative and quantitative abnormalities in myeloid cells leading to both venous/arterial thrombosis and coagulopathy.^20–22^ Similarly, COVID-19 severity is closely related to pro-inflammatory markers,^23^ and is also associated with both thrombosis and coagulopathy.^24,25^ Therefore, patients with myeloproliferative neoplasms may be particularly susceptible to worse COVID-19 outcomes.^26^ To our knowledge, this finding has not been previously described in comparative studies, perhaps because these patients have been excluded or under-represented in cancer cohort studies such as CCC19 and N3C. Some supportive evidence does exist. In a non-comparative study, Salisbury et al.^27^ highlighted a high rate of adverse outcomes in patients with myeloproliferative neoplasms and COVID-19, especially upon ruxolitinib withdrawal. Other groups have reported a similar or decreased risk of mortality in patients with myeloproliferative neoplasms compared to those with other hematologic malignancies.^28–30^ In our study, we did not find that hospitalized patients with myeloproliferative neoplasms had a higher risk of severe COVID-19, but this analysis is limited by the small sample size.

Two antineoplastic medications were found to be associated with an increased risk of hospitalization. Venetoclax, a Bcl2 inhibitor, is commonly used in the treatment of chronic lymphocytic leukemia as a monotherapy, and in combination with other therapies for acute myeloid leukemia. Adverse outcomes have been previously described in patients receiving venetoclax for chronic lymphocytic leukemia in a non-comparative study, but not in any large, comparative study to our knowledge.^31^ Its association with increased risk of hospitalization may be related to the negative effect on immune function via reduced interferon-alpha production and dendritic cells depletion; pneumonia is a known toxicity or venetoclax.^32^ Another potential mechanism involves ACE-2 and bcl-2. Motaghinejad et al.^33^ postulated that increased COVID-19 mortality is partially driven by decreased ACE2 expression in the pulmonary and cardiovascular systems, causing destabilization of Bcl-2 and dysregulation of apoptosis. This dysregulation may be compounded for patients receiving venetoclax, a Bcl-2 inhibitor, leading to cardiopulmonary complications.

Methotrexate was also associated with increased risk of hospitalization. As a cytotoxic chemotherapy, methotrexate may increase susceptibility to COVID-19 complications through myelosuppression. Though most studies suggest that chemotherapy in general is a risk factor for worse COVID-19 outcomes, several studies have not confirmed the association, likely due to the heterogeneity of different chemotherapy agents and regimens.^4,9,34^ Methotrexate as a risk factor has not been studied in cancer patients, but findings for low-dose methotrexate in patients with rheumatologic conditions have been mixed.^35,36^ Despite the biologic rationale for poor COVID-19 outcomes in patients receiving venetoclax or methotrexate, we did not find a consistent association across other outcomes. For example, there was no increased risk of severe COVID-19 in hospitalized patients receiving these therapies. Though this negative finding should be interpreted with caution given the small sample sizes, we cannot exclude the possibility that these observations are coincidental given the high number of individual therapies included. Further confirmatory studies investigating these potential risk factors should be performed.

We also identified known risk factors for hospitalization following COVID-19. These risk factors included older age, Asian race, Hispanic or Latino ethnicity, coronary artery disease, chronic kidney disease, diabetes mellitus, and chronic obstructive pulmonary disease. Numerous prior studies have demonstrated these associations.^5,8,37–39^

This study has several strengths, including one of the largest cancer cohorts to date, a diverse cohort, and use of a novel database. There are several limitations. The database does not contain other risk factors for severe COVID-19, including cancer stage, smoking status, poor performance status, and socioeconomic variables such as insurance status. Similarly, the database did not allow us to discriminate between patients with active versus non-active cancer. There may be selection bias from potential over-representation of patients cared for at academic centers, as well as the inclusion of only patients who underwent SARS-CoV-2 testing. Lastly, we could not ascertain outcomes of patients who sought care outside the UC health system.

### Conclusion

As the COVID-19 pandemic continues in regions without high levels of vaccination and new, highly transmissible variants develop, it is important to remain vigilant of risk factors for severe infection. Close attention will allow us to better prevent and monitor COVID-19 in high-risk patients. Patients with COVID-19 and myeloproliferative neoplasms, and those receiving methotrexate or venetoclax, may be at an increased risk of poor outcomes. Further studies to confirm these associations are needed, as are studies to understand underlying mechanisms. Further investigation is also needed to explain and confirm the lower risk of test positivity in patients with cancer than those without cancer. Lastly, policy makers and health systems should focus on establishing timely, live central databases of electronic health data to provide rapidly accumulating data for future pandemic preparedness, as well as the human capital needed for their maintenance and use.

## Data Availability

Data can be made available upon request, with a data sharing agreement.

## Acknowledgements

We thank Dr. Sharat Israni and Dr. Atul Butte for their support. In addition, this document was prepared as an account of work sponsored by an agency of the United States government. Neither the United States government nor Lawrence Livermore National Security, LLC, nor any of their employees makes any warranty, expressed or implied, or assumes any legal liability or responsibility for the accuracy, completeness, or usefulness of any information, apparatus, product, or process disclosed, or represents that its use would not infringe privately owned rights. Reference herein to any specific commercial product, process, or service by trade name, trademark, manufacturer, or otherwise does not necessarily constitute or imply its endorsement, recommendation, or favoring by the United States government or Lawrence Livermore National Security, LLC. The views and opinions of authors expressed herein do not necessarily state or reflect those of the United States government or Lawrence Livermore National Security, LLC, and shall not be used for advertising or product endorsement purposes.

## References

1. WHO Coronavirus (COVID-19) Dashboard. Accessed September 1, 2021. https://covid19.who.int/

2. Giannakoulis VG, Papoutsi E, Siempos II. Effect of Cancer on Clinical Outcomes of Patients With COVID-19: A Meta-Analysis of Patient Data. JCO Glob Oncol. 2020;6:799–808. doi:10.1200/GO.20.00225

3. Fillmore NR, La J, Szalat RE, et al. Prevalence and Outcome of COVID-19 Infection in Cancer Patients: A National Veterans Affairs Study. J Natl Cancer Inst. 2021;113(6):691–698. doi:10.1093/jnci/djaa159

4. Grivas P, Khaki AR, Wise-Draper TM, et al. Association of clinical factors and recent anticancer therapy with COVID-19 severity among patients with cancer: a report from the COVID-19 and Cancer Consortium. Ann Oncol. 2021;32(6):787–800. doi:10.1016/j.annonc.2021.02.024

5. Harrison SL, Fazio-Eynullayeva E, Lane DA, Underhill P, Lip GYH. Comorbidities associated with mortality in 31,461 adults with COVID-19 in the United States: A federated electronic medical record analysis. PLoS Med. 2020;17(9):e1003321. doi:10.1371/journal.pmed.1003321

6. Rogado J, Obispo B, Pangua C, et al. Covid-19 transmission, outcome and associated risk factors in cancer patients at the first month of the pandemic in a Spanish hospital in Madrid. Clin Transl Oncol. 2020;22(12):2364–2368. doi:10.1007/s12094-020-02381-z

7. Wang Q, Berger NA, Xu R. Analyses of Risk, Racial Disparity, and Outcomes Among US Patients With Cancer and COVID-19 Infection. JAMA Oncol. 2021;7(2):220–227. doi:10.1001/jamaoncol.2020.6178

8. Williamson EJ, Walker AJ, Bhaskaran K, et al. Factors associated with COVID-19-related death using OpenSAFELY. Nature. 2020;584(7821):430–436. doi:10.1038/s41586-020-2521-4

9. Yekedüz E, Utkan G, Ürün Y. A systematic review and meta-analysis: the effect of active cancer treatment on severity of COVID-19. Eur J Cancer. 2020;141:92–104. doi:10.1016/j.ejca.2020.09.028

10. Sharafeldin N, Bates B, Song Q, et al. Outcomes of COVID-19 in Patients With Cancer: Report From the National COVID Cohort Collaborative (N3C). J Clin Oncol. 2021;39(20):2232–2246. doi:10.1200/JCO.21.01074

11. COVID-19 Clinical Data Sets for Research. Published September 17, 2020. Accessed June 17, 2021. https://www.ucbraid.org/cords

12. People with Certain Medical Conditions. Centers for Disease Control and Prevention. Accessed January 1, 2021. https://www.cdc.gov/coronavirus/2019-ncov/need-extra-precautions/people-with-medical-conditions.html

13. van Buuren S, Groothius-Oudshoorn K. mice: Multivariate Imputation by Chained Equations in R. J Stat Softw. 2011;45(3):1–67.

14. Althouse AD. Adjust for Multiple Comparisons? It’s Not That Simple. Ann Thorac Surg. 2016;101(5):1644–1645. doi:10.1016/j.athoracsur.2015.11.024

15. Seabold S, Perktold J. Econometric and statistical modeling with python. In: Proceedings of the 9th Python in Science Conferences. Vol 57.; 2010:61. doi:10.25080/Majora-92bf1922-011

16. Reznikov LR, Norris MH, Vashisht R, et al. Identification of antiviral antihistamines for COVID-19 repurposing. Biochem Biophys Res Commun. Published online December 3, 2020. doi:10.1016/j.bbrc.2020.11.095

17. Goodman KW. Ethics in Health Informatics. Yearb Med Inform. 2020;29(1):26–31. doi:10.1055/s-0040-1701966

18. Bertuzzi AF, Marrari A, Gennaro N, et al. Low Incidence of SARS-CoV-2 in Patients with Solid Tumours on Active Treatment: An Observational Study at a Tertiary Cancer Centre in Lombardy, Italy. Cancers (Basel). 2020;12(9). doi:10.3390/cancers12092352

19. Johannesen TB, Smeland S, Aaserud S, et al. COVID-19 in Cancer Patients, Risk Factors for Disease and Adverse Outcome, a Population-Based Study From Norway. Front Oncol. 2021;11:652535. doi:10.3389/fonc.2021.652535

20. Landolfi R, Rocca B, Patrono C. Bleeding and thrombosis in myeloproliferative disorders: mechanisms and treatment. Crit Rev Oncol Hematol. 1995;20(3):203–222. doi:10.1016/1040-8428(94)00164-O

21. Hermouet S, Bigot-Corbel E, Gardie B. Pathogenesis of Myeloproliferative Neoplasms: Role and Mechanisms of Chronic Inflammation. Mediators Inflamm. 2015;2015:145293. doi:10.1155/2015/145293

22. Brodmann S, Passweg JR, Gratwohl A, Tichelli A, Skoda RC. Myeloproliferative disorders: complications, survival and causes of death. Ann Hematol. 2000;79(6):312–318. doi:10.1007/s002779900136

23. Gong J, Dong H, Xia Q-S, et al. Correlation analysis between disease severity and inflammation-related parameters in patients with COVID-19: a retrospective study. BMC Infect Dis. 2020;20(1):963. doi:10.1186/s12879-020-05681-5

24. Al-Ani F, Chehade S, Lazo-Langner A. Thrombosis risk associated with COVID-19 infection. A scoping review. Thromb Res. 2020;192:152–160. doi:10.1016/j.thromres.2020.05.039

25. Iba T, Levy JH, Levi M, Thachil J. Coagulopathy in COVID-19. J Thromb Haemost. 2020;18(9):2103–2109. doi:10.1111/jth.14975

26. Kamaz B, Mullally A. COVID-19 and myeloproliferative neoplasms: some considerations. Leukemia. 2021;35(1):279–281. doi:10.1038/s41375-020-01070-8

27. Salisbury RA, Curto-Garcia N, O’Sullivan J, et al. Results of a national UK physician reported survey of COVID-19 infection in patients with a myeloproliferative neoplasm. Leukemia. Published online February 12, 2021. doi:10.1038/s41375-021-01143-2

28. Passamonti F, Cattaneo C, Arcaini L, et al. Clinical characteristics and risk factors associated with COVID-19 severity in patients with haematological malignancies in Italy: a retrospective, multicentre, cohort study. Lancet Haematol. 2020;7(10):e737–e745. doi:10.1016/S2352-3026(20)30251-9

29. Yigenoglu TN, Ata N, Altuntas F, et al. The outcome of COVID-19 in patients with hematological malignancy. J Med Virol. 2021;93(2):1099–1104. doi:10.1002/jmv.26404

30. García-Suárez J, de la Cruz J, Cedillo Á, et al. Impact of hematologic malignancy and type of cancer therapy on COVID-19 severity and mortality: lessons from a large population-based registry study. J Hematol Oncol. 2020;13(1):133. doi:10.1186/s13045-020-00970-7

31. Fürstenau M, Langerbeins P, De Silva N, et al. COVID-19 among fit patients with CLL treated with venetoclax-based combinations. Leukemia. 2020;34(8):2225–2229. doi:10.1038/s41375-020-0941-7

32. Reinwald M, Silva JT, Mueller NJ, et al. ESCMID Study Group for Infections in Compromised Hosts (ESGICH) Consensus Document on the safety of targeted and biological therapies: an infectious diseases perspective (Intracellular signaling pathways: tyrosine kinase and mTOR inhibitors). Clin Microbiol Infect. 2018;24 Suppl 2:S53–S70. doi:10.1016/j.cmi.2018.02.009

33. Motaghinejad M, Abbasi Mesrabadi M, Gholami M, Safari S. Possible Involvement of Autophagy or Apoptosis Dysregulation in Infection with COVID-19 Virus as the Main Cause of Mortality: Hypothetical Function of ACE-2/Mas / Ang (1-7) Signaling Pathway and Proposal of a Post-infection Treatment Strategy. Crit Comments Biomed. 2020;1(2):100–124.

34. Jee J, Foote MB, Lumish M, et al. Chemotherapy and COVID-19 Outcomes in Patients With Cancer. J Clin Oncol. 2020;38(30):3538–3546. doi:10.1200/JCO.20.01307

35. Bjornsson AH, Grondal G, Kristjansson M, et al. Prevalence, admission rates and hypoxia due to COVID-19 in patients with rheumatic disorders treated with targeted synthetic or biologic disease modifying antirheumatic drugs or methotrexate: a nationwide study from Iceland. Ann Rheum Dis. Published online January 5, 2021. doi:10.1136/annrheumdis-2020-219564

36. Yousaf A, Gayam S, Feldman S, Zinn Z, Kolodney M. Clinical outcomes of COVID-19 in patients taking tumor necrosis factor inhibitors or methotrexate: A multicenter research network study. J Am Acad Dermatol. 2021;84(1):70–75. doi:10.1016/j.jaad.2020.09.009

37. Sze S, Pan D, Nevill CR, et al. Ethnicity and clinical outcomes in COVID-19: A systematic review and meta-analysis. EClinicalMedicine. 2020;29:100630. doi:10.1016/j.eclinm.2020.100630

38. CDC COVID-19 Response Team. Severe Outcomes Among Patients with Coronavirus Disease 2019 (COVID-19) - United States, February 12-March 16, 2020. MMWR Morb Mortal Wkly Rep. 2020;69(12):343–346. doi:10.15585/mmwr.mm6912e2

39. CDC COVID-19 Response Team. Preliminary Estimates of the Prevalence of Selected Underlying Health Conditions Among Patients with Coronavirus Disease 2019 - United States, February 12-March 28, 2020. MMWR Morb Mortal Wkly Rep. 2020;69(13):382–386. doi:10.15585/mmwr.mm6913e2

