## Supplementary Data for "COVID-19 Outcomes in Patients with Cancer: Findings from the University of California Health System Database"

**S1: Flow diagram illustrating study cohort selection for analyses**

**Cohort 1**
Adult patients undergoing SARS-CoV-2 testing between February 1, 2020 and December 31, 2020
*N*=409,462

Cancer history exclusion

- No clinical encounter associated with a cancer^a^ diagnosis within 1 year prior to first SARS-CoV-2 test date. *N*=407,681

Other exclusion

- Unknown and Other Gender. *N*=464.

**Cohort 2**

Adult patients with COVID-19 and a history of cancer

*N*=1,781

Hospitalization exclusion

- Not hospitalized within 30 days after first SARS-CoV-2 test date. *N*=1,393

**Cohort 3**

Adult patients with a history of cancer who were hospitalized after COVID-19

*N*=388

^a^ Patients with basal and squamous cell cutaneous cancers were excluded.

**S2. Categorization of antineoplastic systemic therapies**

|  | **Antibody** | **Chemo-therapy** | **Hormone therapy** | **Immune-based therapy** | **Tyrosine kinase inhibitor** | **Other cytotoxic therapy** | **Other targeted therapy** |
| --- | --- | --- | --- | --- | --- | --- | --- |
| Anti-EGFR: e.g., cetuximab | x |  |  |  |  |  |  |
| Anti-HER2: e.g., trastuzumab, pertuzumab | x |  |  |  |  |  |  |
| Anti-CD20: e.g., rituximab | x |  |  |  |  |  |  |
| Anti-CD38: e.g., daratumumab | x |  |  |  |  |  |  |
| Anti-angiogenesis: e.g., bevacizumab | x |  |  |  |  |  |  |
| Other: e.g., elotuzumab | x |  |  |  |  |  |  |
| Cytotoxic chemotherapy: e.g., etoposide, methotrexate, temozolamide |  | x |  |  |  |  |  |
| Androgen deprivation therapies: e.g., leuprolide, degarelix, abiraterone. |  |  | x |  |  |  |  |
| Estrogen-targeted therapies: e.g., tamoxifen |  |  | x |  |  |  |  |
| Checkpoint inhibitors: e.g., pembrolizumab |  |  |  | x |  |  |  |
| Cellular therapies: e.g., tisagenlecleucel |  |  |  | x |  |  |  |
| Cancer vaccines: e.g., sipuleucel-T |  |  |  | x |  |  |  |
| Immunostimulants: e.g., interferon-alfa 2b |  |  |  | x |  |  |  |
| VEGF inhibitor: e.g., sunitinib |  |  |  |  | x |  |  |
| BCR/ABL inhibitor: e.g., imatinib |  |  |  |  | x |  |  |
| JAK2 inhibitor: e.g., ruxolitinib, fedratinib |  |  |  |  | x |  |  |
| BTK inhibitor: e.g., ibrutinib |  |  |  |  | x |  |  |
| EGFR inhibitor: e.g., erlotinib |  |  |  |  | x |  |  |
| Other: e.g., vandetanib |  |  |  |  | x |  |  |
| mTOR inhibitor: e.g., everolimus |  |  |  |  |  | x |  |
| CDK4/6 inhibitor: e.g., palbociclib |  |  |  |  |  | x |  |
| Other: e.g., lenalidomide |  |  |  |  |  | x |  |
| HDAC inhibitor: e.g., belinostat, romidepsin |  |  |  |  |  | x |  |
| BCL2 inhibitor: e.g., venetoclax |  |  |  |  |  |  | x |
| PARP inhibitor: e.g., olaparib |  |  |  |  |  |  | x |
| Antibody-drug conjugate: e.g., enfortumab vedotin |  |  |  |  |  |  | x |
| PI3K inhibitor: e.g., copanlisib |  |  |  |  |  |  | x |
| BRAF/MEK inhibitor: e.g., dabrafenib |  |  |  |  |  |  | x |
| Other: e.g., enasdenib |  |  |  |  |  |  | x |

**S3. Antineoplastic systemic therapies received by 1,781 cancer patients undergoing SARS-CoV-2 testing**

| **Therapy** | **Category** | ***N* (%)** |
| --- | --- | --- |
| Androgen deprivation therapies (e.g.,  leuprolide acetate, degarelix, abiraterone etc.) | Hormone | 42 (2.31) |
| Arsenic | Chemotherapy | 1 (0.06) |
| Bevacizumab | Antibody | 13 (0.72) |
| Blinatumomab | Antibody | 2 (0.11) |
| Cladribine | Chemotherapy | 1 (0.06) |
| Clofarabine | Chemotherapy | 1 (0.06) |
| Cyclophosphamide | Chemotherapy | 2 (0.11) |
| Cytarabine | Chemotherapy | 1 (0.06) |
| Daratumumab | Antibody | 4 (0.22) |
| Dasatinib | TKI | 7 (0.39) |
| Decitabine | Chemotherapy | 1 (0.06) |
| Docetaxel | Chemotherapy | 9 (0.50) |
| Doxorubicin | Chemotherapy | 20 (1.10) |
| Encorafenib | Other targeted | 2 (0.11) |
| Entrectinib | Other targeted | 1 (0.06) |
| Eribulin | Chemotherapy | 1 (0.06) |
| Etoposide | Chemotherapy | 20 (1.10) |
| Everolimus | Other cytotoxic | 6 (0.33) |
| Exemestane | Hormone | 11 (0.61) |
| Fedratinib | TKI | 3 (0.17) |
| Gemcitabine | Chemotherapy | 4 (0.22) |
| Ifosfamide | Chemotherapy | 6 (0.33) |
| Imatinib | TKI | 4 (0.22) |
| Lenalidomide | Other cytotoxic | 20 (1.10) |
| Letrozole | Hormone | 28 (1.54) |
| Lomustine | Chemotherapy | 2 (0.11) |
| Methotrexate | Chemotherapy | 24 (1.32) |
| Mitomycin | Chemotherapy | 1 (0.06) |
| Mitoxantrone | Chemotherapy | 1 (0.06) |
| Neratinib | Other cytotoxic | 1 (0.06) |
| Olaparib | Other targeted | 1 (0.06) |
| Osimertinib | TKI | 3 (0.17) |
| Oxaliplatin | Chemotherapy | 19 (1.05) |
| Paclitaxel | Chemotherapy | 42 (2.31) |
| Palbociclib | Other cytotoxic | 2 (0.11) |
| Panitumumab | Antibody | 6 (0.33) |
| Pemetrexed | Chemotherapy | 8 (0.44) |
| Pertuzumab | Antibody | 10 (0.55) |
| Pomalidomide | Other cytotoxic | 4 (0.22) |
| Ramucirumab | Antibody | 3 (0.17) |
| Regorafenib | TKI | 1 (0.06) |
| Romidepsin | Other cytotoxic | 1 (0.06) |
| Ruxolitinib | TKI | 7 (0.39) |
| Sirolimus | Other cytotoxic | 8 (0.44) |
| Tamoxifen | Hormone | 21 (1.16) |
| Temozolamide | Chemotherapy | 7 (0.39) |
| Thiotepa | Chemotherapy | 1 (0.06) |
| Trastuzumab | Antibody | 11 (0.61) |
| Venetoclax | Other targeted | 14 (0.77) |
| Vinca alkaloid | Chemotherapy | 35 (1.93) |
| Vismodegib | Other targeted | 1 (0.06) |

TKI = Tyrosine Kinase Inhibitor

**S4: Risk of SARS-CoV-2 test positivity among 409,926 adult patients undergoing testing, with cancer types delineated**

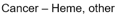

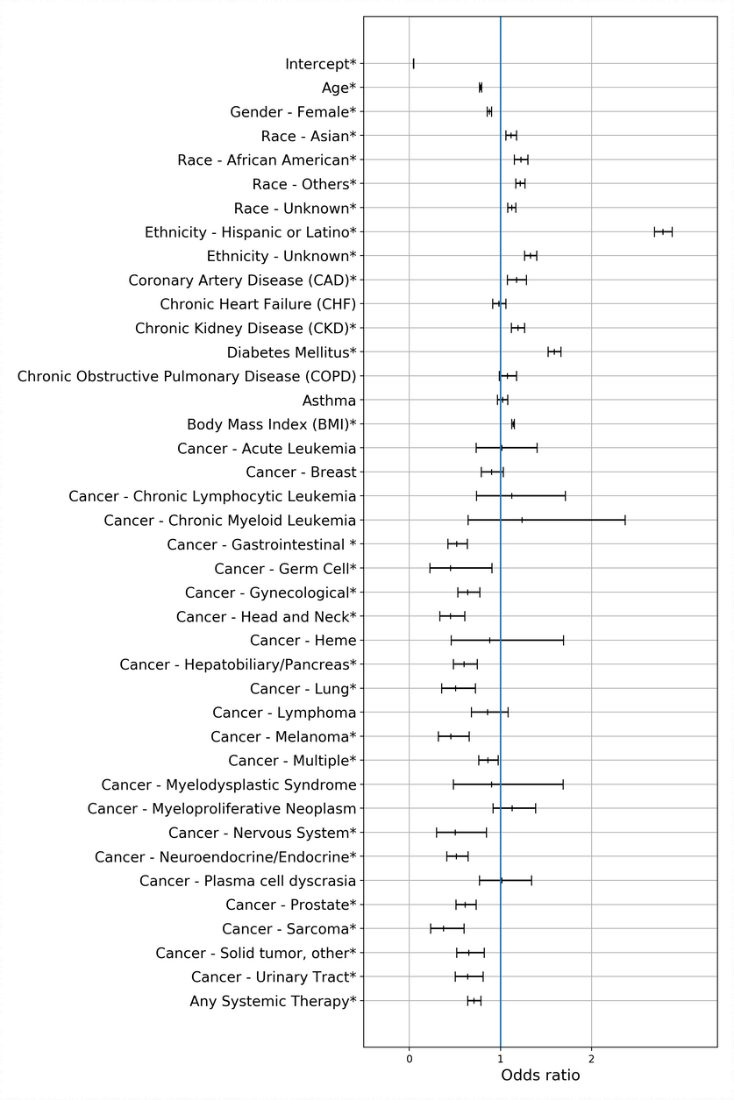

**S5: Risk of 30-day intensive care, mechanical ventilation, or death among 388 adult cancer patients with a history of cancer and positive SARS-CoV-2, with interaction term**

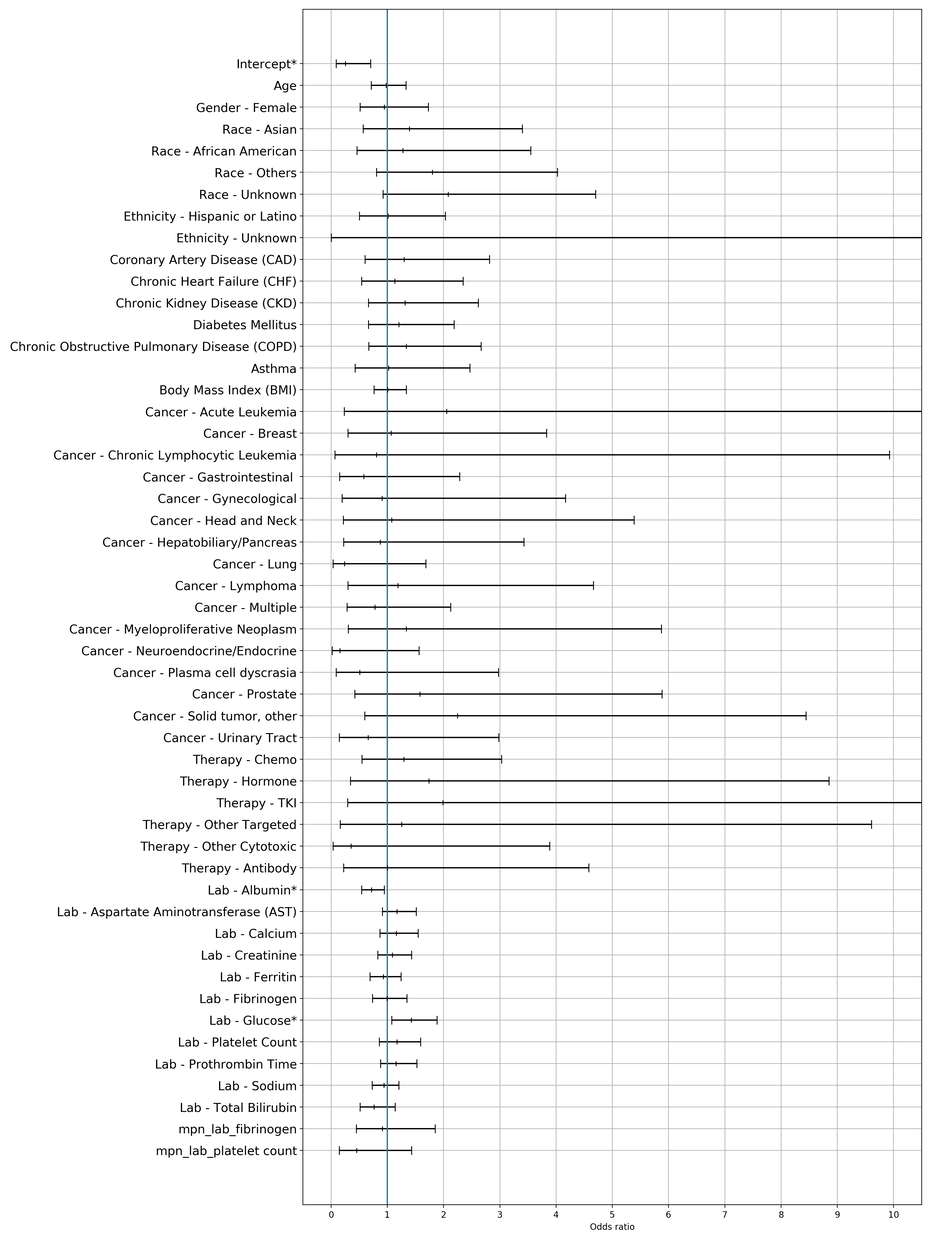

**S6: Risk of SARS-CoV-2 test positivity among 409,462 adult patients undergoing testing**

|  | **Adjusted odds ratio** | **95% CI lower bound** | **95% CI upper bound** | ***P*-value** |
| --- | --- | --- | --- | --- |
| **Age** | 0.78 | 0.77 | 0.79 | <0.001 |
| **Gender** |  | | | |
| Male | Reference | | | |
| Female | 0.88 | 0.86 | 0.90 | <0.001 |
| **Race** |  | | | |
| White | Reference | | | |
| Asian | 1.11 | 1.06 | 1.17 | <0.001 |
| Black or African American | 1.22 | 1.15 | 1.30 | <0.001 |
| Other | 1.21 | 1.16 | 1.26 | <0.001 |
| Unknown | 1.12 | 1.08 | 1.17 | <0.001 |
| **Ethnicity** |  | | | |
| Not Hispanic or Latino | Reference | | | |
| Hispanic or Latino | 2.78 | 2.68 | 2.87 | <0.001 |
| Unknown | 1.32 | 1.26 | 1.39 | <0.001 |
| **Comorbidities** |  |  |  |  |
| Coronary Artery Disease | 1.16 | 1.07 | 1.27 | 0.001 |
| Congestive Heart Failure | 0.99 | 0.92 | 1.07 | 0.815 |
| Chronic Kidney Disease | 1.20 | 1.13 | 1.27 | <0.001 |
| Diabetes Mellitus | 1.59 | 1.52 | 1.66 | <0.001 |
| Chronic Obstructive Pulmonary Disease | 1.07 | 0.98 | 1.17 | 0.126 |
| Asthma | 1.02 | 0.97 | 1.08 | 0.467 |
| Body Mass Index | 1.14 | 1.12 | 1.15 | <0.001 |
| **History of cancer** | 0.69 | 0.66 | 0.73 | <0.001 |
| **Any systemic therapy** | 0.76 | 0.69 | 0.84 | <0.001 |

**S7: Risk of 30-day hospitalization following a positive SARS-CoV-2 test among 1,781 adult cancer patients**

|  | **Adjusted odds ratio** | **95% CI lower bound** | **95% CI upper bound** | ***P*-value** |
| --- | --- | --- | --- | --- |
| **Age** | 1.34 | 1.15 | 1.57 | <0.001 |
| **Gender** |  | | | |
| Male | Reference | | | |
| Female | 0.78 | 0.58 | 1.03 | 0.083 |
| **Race** |  | | | |
| White | Reference | | | |
| Asian | 1.92 | 1.23 | 2.98 | 0.004 |
| Black or African American | 1.17 | 0.69 | 1.97 | 0.560 |
| Other | 0.87 | 0.59 | 1.28 | 0.474 |
| Unknown | 0.77 | 0.52 | 1.15 | 0.203 |
| **Ethnicity** |  | | | |
| Not Hispanic or Latino | Reference | | | |
| Hispanic or Latino | 1.96 | 1.41 | 2.73 | <0.001 |
| Unknown | 0.39 | 0.09 | 1.71 | 0.211 |
| **Comorbidities** |  |  |  |  |
| Coronary Artery Disease | 1.76 | 1.15 | 2.70 | 0.009 |
| Congestive Heart Failure | 1.11 | 0.76 | 1.64 | 0.588 |
| Chronic Kidney Disease | 1.52 | 1.08 | 2.14 | 0.016 |
| Diabetes Mellitus | 1.65 | 1.25 | 2.18 | <0.001 |
| Chronic Obstructive Pulmonary Disease | 3.12 | 2.07 | 4.72 | <0.001 |
| Asthma | 0.96 | 0.63 | 1.48 | 0.860 |
| Body Mass Index | 0.99 | 0.87 | 1.12 | 0.819 |
| **Cancer type** |  | | | |
| Unspecified | Reference | | | |
| Acute leukemia | 0.57 | 0.19 | 1.71 | 0.318 |
| Breast | 0.82 | 0.42 | 1.61 | 0.569 |
| Chronic lymphocytic leukemia | 0.93 | 0.29 | 2.96 | 0.900 |
| Chronic myeloid leukemia | 0.00 | 0.00 | >99 | 0.887 |
| Gastrointestinal | 1.21 | 0.60 | 2.45 | 0.588 |
| Germ cell | 1.70 | 0.18 | 16.41 | 0.644 |
| Gynecological | 0.82 | 0.39 | 1.77 | 0.620 |
| Head and neck | 1.40 | 0.58 | 3.38 | 0.450 |
| Hematologic, other | 1.27 | 0.23 | 6.97 | 0.784 |
| Hepatobiliary/Pancreas | 1.25 | 0.61 | 2.57 | 0.538 |
| Lung | 1.75 | 0.68 | 4.52 | 0.248 |
| Lymphoma | 1.56 | 0.74 | 3.27 | 0.240 |
| Melanoma | 0.13 | 0.02 | 1.13 | 0.064 |
| Multiple | 1.49 | 0.84 | 2.64 | 0.173 |
| Myelodysplastic syndrome | 0.73 | 0.13 | 4.22 | 0.726 |
| Myeloproliferative neoplasm | 2.51 | 1.29 | 4.89 | 0.007 |
| Nervous system | 1.83 | 0.44 | 7.53 | 0.404 |
| Neuroendocrine/ Endocrine | 0.89 | 0.39 | 2.06 | 0.791 |
| Plasma cell dyscrasia | 1.70 | 0.74 | 3.92 | 0.212 |
| Prostate | 0.95 | 0.47 | 1.91 | 0.884 |
| Sarcoma | 1.15 | 0.23 | 5.66 | 0.865 |
| Solid tumor, other | 1.43 | 0.67 | 3.04 | 0.355 |
| Urinary tract | 1.04 | 0.47 | 2.30 | 0.923 |
| **Cancer therapy type** |  |  |  |  |
| Chemotherapy | 1.46 | 0.95 | 2.26 | 0.086 |
| Hormone | 0.78 | 0.36 | 1.69 | 0.536 |
| Tyrosine kinase inhibitor | 1.74 | 0.59 | 5.09 | 0.314 |
| Other targeted | 3.64 | 1.21 | 10.98 | 0.022 |
| Other cytotoxic | 1.05 | 0.45 | 2.47 | 0.911 |
| Antibody | 1.59 | 0.73 | 3.42 | 0.241 |

**S8: Risk of 30-day hospitalization following a positive SARS-CoV-2 test among 1,781 adult cancer patients, individual therapies delineated**

|  | **Adjusted odds ratio** | **95% CI lower bound** | **95% CI upper bound** | ***P*-value** |
| --- | --- | --- | --- | --- |
| **Age** | 1.34 | 1.15 | 1.57 | <0.001 |
| **Gender** |  | | | |
| Male | Reference | | | |
| Female | 0.79 | 0.59 | 1.06 | 0.119 |
| **Race** |  | | | |
| White | Reference | | | |
| Asian | 1.92 | 1.23 | 3.00 | 0.004 |
| Black or African American | 1.15 | 0.68 | 1.95 | 0.601 |
| Other | 0.89 | 0.60 | 1.32 | 0.572 |
| Unknown | 0.73 | 0.49 | 1.10 | 0.136 |
| **Ethnicity** |  | | | |
| Not Hispanic or Latino | Reference | | | |
| Hispanic or Latino | 1.94 | 1.39 | 2.71 | <0.001 |
| Unknown | 0.40 | 0.09 | 1.75 | 0.221 |
| **Comorbidities** |  |  |  |  |
| Coronary Artery Disease | 1.74 | 1.13 | 2.67 | 0.012 |
| Congestive Heart Failure | 1.16 | 0.78 | 1.73 | 0.454 |
| Chronic Kidney Disease | 1.49 | 1.05 | 2.11 | 0.026 |
| Diabetes Mellitus | 1.72 | 1.30 | 2.29 | <0.001 |
| Chronic Obstructive Pulmonary Disease | 3.04 | 2.00 | 4.63 | <0.001 |
| Asthma | 0.97 | 0.63 | 1.50 | 0.882 |
| Body Mass Index | 0.98 | 0.86 | 1.12 | 0.821 |
| **Cancer type** |  | | | |
| Unspecified | Reference | | | |
| Acute leukemia | 0.46 | 0.14 | 1.58 | 0.219 |
| Breast | 0.86 | 0.43 | 1.72 | 0.673 |
| Chronic lymphocytic leukemia | 0.92 | 0.29 | 2.96 | 0.895 |
| Chronic myeloid leukemia | 0.00 | 0.00 | >99 | 0.962 |
| Gastrointestinal | 1.23 | 0.60 | 2.52 | 0.577 |
| Germ cell | 1.74 | 0.18 | 16.68 | 0.632 |
| Gynecological | 0.86 | 0.40 | 1.85 | 0.707 |
| Head and neck | 1.42 | 0.59 | 3.42 | 0.435 |
| Hematologic, other | 1.25 | 0.23 | 6.90 | 0.795 |
| Hepatobiliary/Pancreas | 1.28 | 0.62 | 2.63 | 0.505 |
| Lung | 1.94 | 0.75 | 5.03 | 0.175 |
| Lymphoma | 1.50 | 0.69 | 3.23 | 0.306 |
| Melanoma | 0.13 | 0.02 | 1.16 | 0.068 |
| Multiple | 1.57 | 0.88 | 2.79 | 0.123 |
| Myelodysplastic syndrome | 0.94 | 0.15 | 5.87 | 0.951 |
| Myeloproliferative neoplasm | 2.43 | 1.24 | 4.75 | 0.010 |
| Nervous system | 1.65 | 0.37 | 7.26 | 0.509 |
| Neuroendocrine/ Endocrine | 0.89 | 0.39 | 2.06 | 0.787 |
| Plasma cell dyscrasia | 1.52 | 0.64 | 3.61 | 0.347 |
| Prostate | 0.97 | 0.48 | 1.96 | 0.931 |
| Sarcoma | 1.25 | 0.25 | 6.20 | 0.785 |
| Solid tumor, other | 1.36 | 0.63 | 2.92 | 0.437 |
| Urinary tract | 1.06 | 0.48 | 2.34 | 0.890 |
| **Individual medication** |  | | | |
| Androgen deprivation therapy | 0.56 | 0.17 | 1.89 | 0.349 |
| Bevacizumab | 1.54 | 0.37 | 6.35 | 0.554 |
| Dasatinib | 0.00 | 0.00 | >99 | 0.980 |
| Docetaxel | 2.47 | 0.54 | 11.36 | 0.245 |
| Doxorubicin | 0.75 | 0.18 | 3.16 | 0.697 |
| Etoposide | 1.28 | 0.34 | 4.85 | 0.714 |
| Everolimus | 3.38 | 0.48 | 24.05 | 0.223 |
| Exemestane | 0.00 | 0.00 | >99 | 0.984 |
| Ifosfamide | 3.47 | 0.51 | 23.76 | 0.205 |
| Lenalidomide | 2.01 | 0.66 | 6.11 | 0.220 |
| Letrozole | 0.65 | 0.14 | 2.95 | 0.578 |
| Methotrexate | 3.65 | 1.17 | 11.37 | 0.026 |
| Oxaliplatin | 2.38 | 0.86 | 6.59 | 0.094 |
| Paclitaxel | 0.44 | 0.14 | 1.36 | 0.154 |
| Panitumumab | 0.53 | 0.05 | 5.53 | 0.598 |
| Pemetrexed | 0.35 | 0.05 | 2.32 | 0.275 |
| Pertuzumab | 4.09 | 0.38 | 43.82 | 0.244 |
| Ruxolitinib | >99 | 0.00 | >99 | 0.973 |
| Sirolimus | 0.00 | 0.00 | >99 | 0.967 |
| Tamoxifen | 1.54 | 0.41 | 5.78 | 0.523 |
| Temozolamide | 3.47 | 0.54 | 22.26 | 0.189 |
| Trastuzumab | 0.87 | 0.09 | 8.81 | 0.906 |
| Venetoclax | 3.63 | 1.02 | 12.92 | 0.046 |
| Vinca alkaloid | 1.22 | 0.41 | 3.68 | 0.720 |

**S9: Risk of 30-day intensive care, mechanical ventilation, or death among 388 adult cancer patients with a history of cancer and positive SARS-CoV-2**

|  | **Adjusted odds ratio** | **95% CI lower bound** | **95% CI upper bound** | ***P*-value** |
| --- | --- | --- | --- | --- |
| **Age** | 0.98 | 0.72 | 1.34 | 0.906 |
| **Gender** |  | | | |
| Male | Reference | | | |
| Female | 1.01 | 0.56 | 1.85 | 0.962 |
| **Race** |  | | | |
| White | Reference | | | |
| Asian | 1.41 | 0.58 | 3.43 | 0.447 |
| Black or African American | 1.31 | 0.47 | 3.65 | 0.606 |
| Other | 1.75 | 0.79 | 3.88 | 0.171 |
| Unknown | 2.09 | 0.93 | 4.72 | 0.075 |
| **Ethnicity** |  | | | |
| Not Hispanic or Latino | Reference | | | |
| Hispanic or Latino | 1.03 | 0.51 | 2.07 | 0.929 |
| Unknown | >99 | 0.00 | >99 | 0.916 |
| **Comorbidities** |  | | | |
| Coronary Artery Disease | 1.23 | 0.57 | 2.64 | 0.604 |
| Congestive Heart Failure | 1.11 | 0.54 | 2.31 | 0.772 |
| Chronic Kidney Disease | 1.31 | 0.66 | 2.58 | 0.442 |
| Diabetes Mellitus | 1.19 | 0.66 | 2.16 | 0.558 |
| Chronic Obstructive Pulmonary Disease | 1.31 | 0.66 | 2.61 | 0.438 |
| Asthma | 1.04 | 0.43 | 2.51 | 0.932 |
| Body Mass Index | 1.01 | 0.76 | 1.33 | 0.962 |
| **Cancer type** |  | | | |
| Unspecified | Reference | | | |
| Acute leukemia | 2.10 | 0.24 | 18.23 | 0.502 |
| Breast | 1.03 | 0.29 | 3.68 | 0.964 |
| Chronic lymphocytic leukemia | 0.86 | 0.07 | 10.18 | 0.903 |
| Gastrointestinal | 0.58 | 0.15 | 2.26 | 0.431 |
| Gynecological | 0.91 | 0.20 | 4.19 | 0.908 |
| Head and neck | 1.13 | 0.23 | 5.65 | 0.884 |
| Hepatobiliary/Pancreas | 0.84 | 0.21 | 3.31 | 0.807 |
| Lung | 0.25 | 0.04 | 1.78 | 0.167 |
| Lymphoma | 1.16 | 0.29 | 4.54 | 0.836 |
| Multiple | 0.79 | 0.29 | 2.14 | 0.637 |
| Myeloproliferative neoplasm | 0.70 | 0.20 | 2.48 | 0.582 |
| Neuroendocrine/ Endocrine | 0.17 | 0.02 | 1.68 | 0.129 |
| Plasma cell dyscrasia | 0.51 | 0.09 | 2.99 | 0.456 |
| Prostate | 1.61 | 0.43 | 6.01 | 0.475 |
| Solid tumor, other | 2.24 | 0.60 | 8.44 | 0.232 |
| Urinary tract | 0.65 | 0.14 | 2.96 | 0.580 |
| **Cancer therapy type** |  | | | |
| Chemotherapy | 1.29 | 0.55 | 3.04 | 0.553 |
| Hormone | 1.70 | 0.34 | 8.61 | 0.519 |
| Tyrosine kinase inhibitor | 2.09 | 0.33 | 13.48 | 0.436 |
| Other targeted | 1.17 | 0.15 | 8.94 | 0.883 |
| Other cytotoxic | 0.33 | 0.03 | 3.65 | 0.369 |
| Antibody | 1.01 | 0.22 | 4.60 | 0.990 |
| **Laboratory test** |  |  |  |  |
| Albumin | 0.73 | 0.56 | 0.96 | 0.027 |
| AST | 1.17 | 0.91 | 1.50 | 0.225 |
| Calcium | 1.19 | 0.89 | 1.58 | 0.238 |
| Creatinine | 1.09 | 0.83 | 1.43 | 0.555 |
| Ferritin | 0.92 | 0.68 | 1.23 | 0.578 |
| Fibrinogen | 0.98 | 0.75 | 1.30 | 0.908 |
| Glucose | 1.45 | 1.10 | 1.90 | 0.008 |
| Platelet count | 1.06 | 0.80 | 1.41 | 0.669 |
| Prothrombin time | 1.19 | 0.92 | 1.55 | 0.178 |
| Sodium | 0.93 | 0.72 | 1.19 | 0.548 |
| Total bilirubin | 0.76 | 0.51 | 1.14 | 0.188 |
